# Subchondral bone length in knee osteoarthritis: A deep learning derived imaging measure and its association with radiographic and clinical outcomes

**DOI:** 10.1101/2021.04.28.21256271

**Authors:** Gary H. Chang, Lisa K. Park, Nina A. Le, Ray S. Jhun, Tejus Surendran, Joseph Lai, Hojoon Seo, Nuwapa Promchotichai, Grace Yoon, Jonathan Scalera, Terence D. Capellini, David T. Felson, Vijaya B. Kolachalama

**Affiliations:** Section of Computational Biomedicine, Department of Medicine, Boston University School of Medicine, Boston, MA, USA – 02118; Department of Radiology, Boston University School of Medicine, Boston, MA, USA – 02118; Department of Human Evolutionary Biology, Harvard University, Cambridge, MA, USA – 02138; Broad Institute of MIT and Harvard, Cambridge, MA, USA 02142; Section of Rheumatology, Department of Medicine, Boston University School of Medicine, Boston, MA, USA – 02118; Centre for Epidemiology, University of Manchester and the NIHR Manchester BRC, Manchester University, NHS Trust, Manchester, UK; Department of Computer Science and Faculty of Computing & Data Sciences, Boston University, Boston, MA, USA – 02215

**Keywords:** Knee Osteoarthritis, Joint Space Narrowing, Knee Replacement, Magnetic Resonance Imaging, Deep Learning

## Abstract

**Objective:** Develop a bone shape measure that reflects the extent of cartilage loss and bone flattening in knee osteoarthritis (OA) and test it against estimates of disease severity.

**Methods:** A fast region-based convolutional neural network was trained to crop the knee joints in sagittal dual-echo steady state MRI sequences obtained from the Osteoarthritis Initiative (OAI). Publicly available annotations of the cartilage and menisci were used as references to annotate the tibia and the femur in 61 knees. Another deep neural network (U-Net) was developed to learn these annotations. Model predictions were compared with radiologist-driven annotations on an independent test set (27 knees). The U-Net was applied to automatically extract the knee joint structures on the larger OAI dataset (9,434 knees). We defined subchondral bone length (SBL), a novel shape measure characterizing the extent of overlying cartilage and bone flattening, and examined its relationship with radiographic joint space narrowing (JSN), concurrent WOMAC pain and disability as well as subsequent partial or total knee replacement (KR). Odds ratios for each outcome were estimated using relative changes in SBL on the OAI dataset into quartiles.

**Result:** Mean SBL values for knees with JSN were consistently different from knees without JSN. Greater changes of SBL from baseline were associated with greater pain and disability. For knees with medial or lateral JSN, the odds ratios between lowest and highest quartiles corresponding to SBL changes for future KR were 5.68 (95% CI:[3.90,8.27]) and 7.19 (95% CI:[3.71,13.95]), respectively.

**Conclusion:** SBL quantified OA status based on JSN severity. It has promise as an imaging marker in predicting clinical and structural OA outcomes.

## INTRODUCTION

Knee osteoarthritis (OA) is one of the most common debilitating conditions among older adults [1], with a burden that is expected to increase around the globe due to factors such as increasing rates of obesity [2]. Because there is no effective disease modifying therapy for knee OA, current clinical management relies in part on identification of radiographic abnormalities such as joint space narrowing (JSN) to assist in diagnosing OA and/or estimate likelihood of progression to severe OA [3-6]. Since it is important to accurately evaluate OA pathophysiology, there is a need to develop additional imaging markers, and in some cases, improve the measurement sensitivity and specificity of existing imaging markers to better predict disease progression and potential clinical outcomes.

Magnetic resonance imaging (MRI) can allow us to visually differentiate joint tissues, which facilitates accurate characterization of several imaging markers of knee OA [7]. Previously, a case-control study on a subset of data from the Osteoarthritis Initiative (OAI) showed that bone shape derived from knee MR images predicted later onset of radiographic knee OA [8]. In other studies, researchers used MRI to identify increased tibial plateau size and subchondral bone attrition during pre-radiographic OA stage [9], and also found variability in knee articular surface geometry in individuals with and without OA [10]. Most of these studies were performed on a small subset of individuals. While findings from these important investigations established a proof-of-principle, it is not trivial to extend such studies to a larger cohort due to the sheer volume of cases and the manual labor needed to precisely annotate different anatomic structures in the knee. Recently, Bowes and colleagues proposed a machine learning-driven measure of femur bone shape (defined as B-score), on a large set of individuals from the OAI cohort (n=4,796) [11]. They showed that B-score was associated with risk of current and future pain, functional limitation and total knee replacement. Few other studies have also attempted to characterize bone shapes in OA [8, 12]. The major contributor to the bone shape metric has been the crust of osteophytes that arises at the margin of the cartilage plate in advanced OA. These marginal osteophytes are probably not a source of pain [13], and their size and number are poor proxies for the severity of nearby cartilage loss [14], which is the signature pathologic feature of OA. We developed a bone shape measure defined as subchondral bone length (SBL), which did not include these marginal osteophytes, but rather was driven by the extent of overlying cartilage, decreasing with cartilage denudation. Bone also flattens with increasing disease severity, increasing its area [15, 16]. We hypothesized that SBL variations increase with the severity of radiographic knee OA, as it accounts for the dynamic changes that occur within the subchondral region due to cartilage loss and bone flattening. We then tested whether SBL correlated with severity of knee OA defined by JSN, concurrent knee pain and disability as well as subsequent partial or total knee replacement.

## METHODS

### Study population

Data were obtained from the Osteoarthritis Initiative (OAI), an NIH funded observational study of persons with or at-risk of knee OA. Study subjects included men and women ranging in age from 45-79 with or at-risk for symptomatic tibiofemoral knee OA (**Table 1**). Minorities made up 20.9% of the population of whom 87.3% were African Americans, 4.5% Asians, and 8.2% fell into the “Other” category. Subjects with contraindications for 3T MRI (such as inflammatory arthritis) were excluded from the OAI study (**Table 1**). The majority of knees at baseline had radiographic severity of KL grades 0-3, with KL grade 4 making up ∼3% of the population. Note that knees with information regarding KL grade also had data regarding JSN grade. All the cases which had both KL and JSN grades were included in the study. The severity of JSN was based on centrally read scores ranging from 0 to 3 (0=normal, 1=mild, 2=moderate and 3=severe) by compartments [17].

**Table 1:**
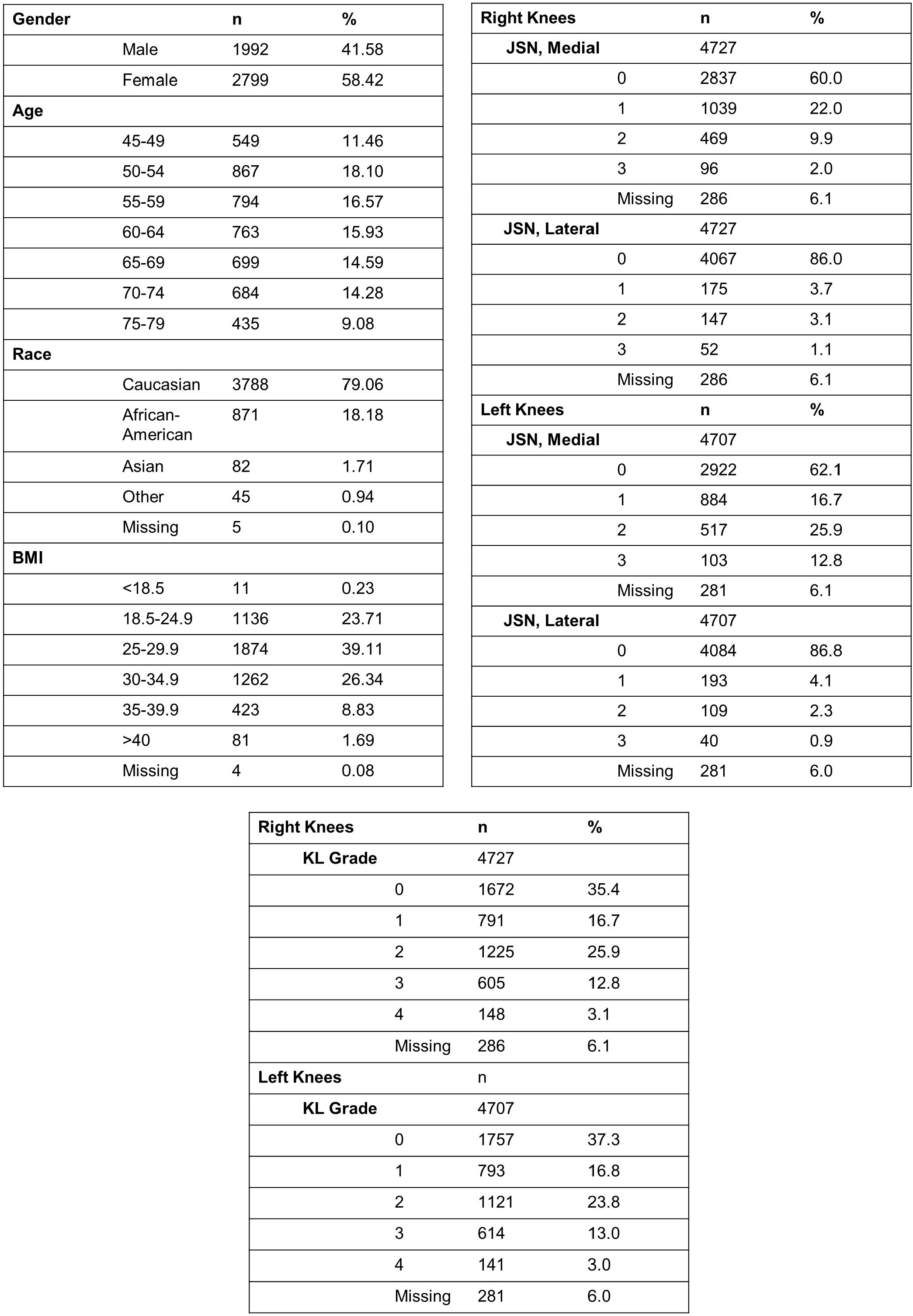
Study population. Cases used from the Osteoarthritis Initiative (OAI) for this study. The table shows the breakdown of participants in terms of their age, race, body mass index, JSN grade and KL grade.

### MRI acquisition and measurements

Three-dimensional (3D) dual-echo in steady state (DESS) sagittal MR sequences of the left and right knees were available on the OAI dataset at baseline (n=9,434 knees). All the scans along with the subject-level baseline clinical data are available on the NIH website (https://nda.nih.gov/oai/), and will be forwarded upon request. We also obtained detailed segmentation masks of the femur cartilage, lateral tibia cartilage and medial tibia cartilage for a small portion of knees from the OAI database (n=88), which were used to train and validate the model. The cartilage and the meniscal image masks were provided by the OA Biomarkers Consortium FNIH Project and performed by iMorphics (Manchester, UK). As per the OAI documentation (https://nda.nih.gov/oai/study_documentation.html), the knees were chosen to represent the OAI database population (primarily moderate and severe KL grades, Male = 45, Female = 43) and should be applicable for the validation process of our study.

The DESS sequence images provide detailed definition of 3D structures and their shapes, particularly of cartilage morphology [18, 19]. The imaging was performed with a 3.0 Tesla magnet using imaging sequence with TR/TE of 16.3/4.7 ms. We selected knees (n=4,727 right and 4,707 left knees) from the OAI baseline exam with DESS sequence MR images. The DESS sequence provides high in-plane and out-of-plane resolution (0.7 mm × 0.365 mm × 0.456 mm) in a time efficient manner. The images encompassed the cartilage of the knee joint as well as the subchondral structures of the tibia and the femur.

### Image preprocessing and a fast region-based convolutional neural network

Image registration and quality check were performed on all the knees as previously described [20]. To focus the network on the joint area, a large-sized bounding box with a dimension of 272×240 pixels was created to identify regions of interest (ROI) comprised of all the subchondral compartments containing the manually annotated femoral and tibia cartilages. The coordinates of the ROI of each knee were subsequently used to train a fast region-based convolutional neural network (Fast R-CNN) model [21] to automatically detect the region of the knee joint (**Figure S1**). The sagittal MRI slices subsequently underwent histogram equalization and their intensity values were normalized to a range of 0 to 1. These slices were then used as the inputs to the neural network used to extract the knee joint structures. More details are available in the supplement.

### Image annotation pipeline

From the OAI database, 88 knee MRIs had expert-driven annotations of the cartilage. These MRIs were chosen as our primary dataset for training and validating the model for both automated segmentation of cartilage and bone shape (**Figure S2**). The 88 knee MRIs were randomly split in the ratio of 7:3 for training and validation, respectively. The 61 knee images for training (n=1,753 2D sagittal slices) were passed through an image-processing pipeline based on Distance Regularized Level Set Evolution (DRLSE) to extract the bone shapes (**Figure S3**). DRLSE is an edge-based active contour model that uses the gradient information of the images to expand the segmented area until it meets the boundary [22]. Once the active contour model was applied, the obtained bone shapes were further manually verified and adjusted to exclude erroneously captured soft tissues in the bone shapes. Finally, the expert-driven segmented regions of cartilage and meniscal areas were superimposed to adjust the bone shapes, where needed. The modified bone shapes from the 61 cases were then used to train a deep neural network (See below and the supplement). The remaining cases (794 2D sagittal slices from 27 knees) were used for model validation and underwent expert-assisted manual annotation of the bone shapes as detailed below.

### Annotation of the knee structures

A board-certified radiologist (JS) manually annotated bone shapes of the knee on the 27 test cases. Using a stylus on a touch screen device, the expert outlined the cortical surface of the tibia and the femur. We traced the SBLs on both the tibia and the femur (direction shown in dotted yellow lines in **Figure 1A**), to capture the shape of the cortical bone of the femur or the tibia in contact with the cartilage.

**Figure 1:**
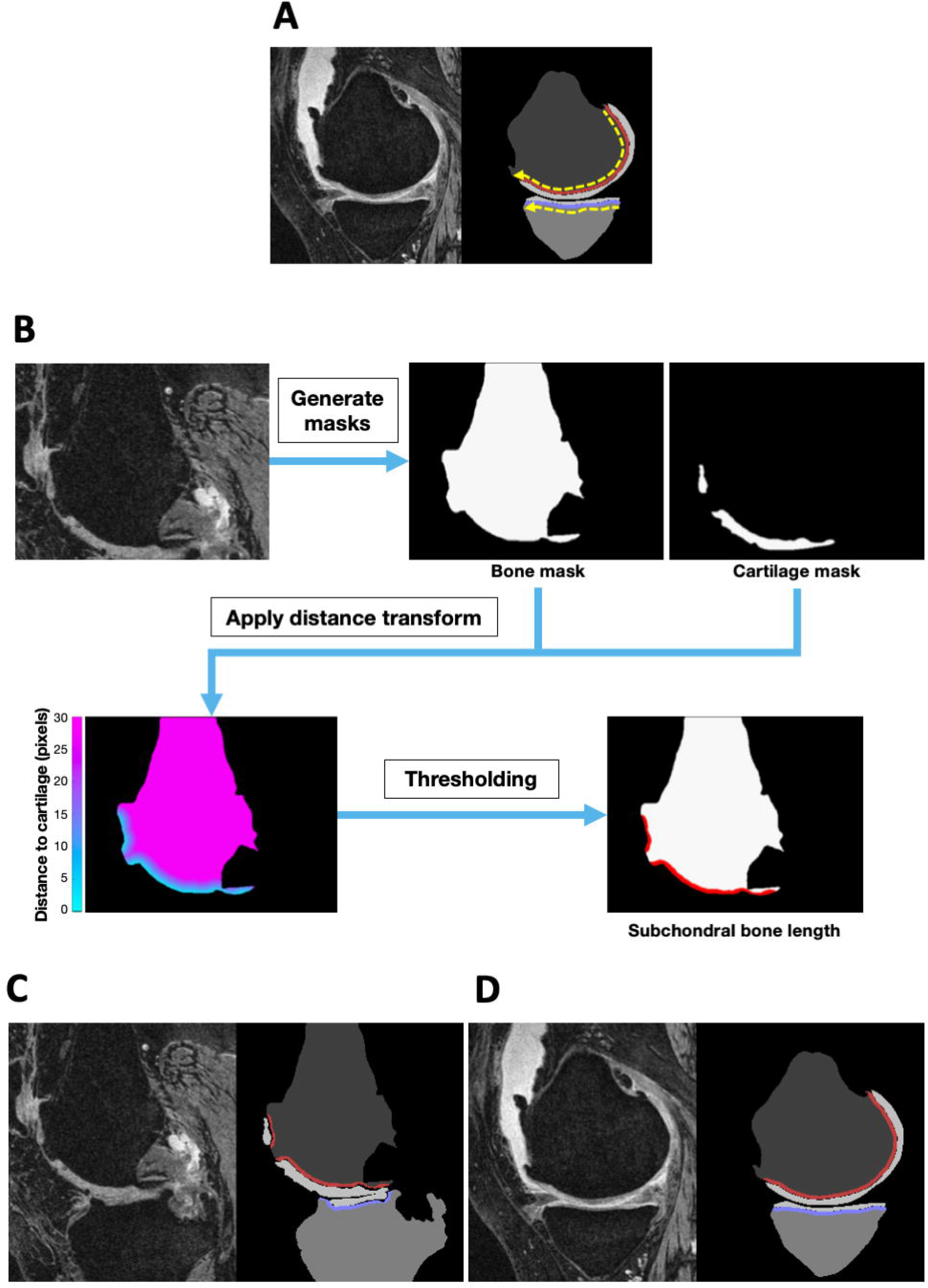
Subchondral bone length. (A) An MRI slice is shown in the sagittal plane along with the traced subchondral bone lengths (SBLs) on both the femur (red line) and the tibia (blue line). Two dotted yellow lines show the direction of tracing to obtain the SBLs on the femur and the tibia, respectively. (B) Estimation of SBL by performing a distance transform using the masks of the bone and the cartilage. (C-D) Examples of the original MRI slices (sagittal view) along with the respective masks generated by the deep learning model. Note the exclusion of the anterior marginal osteophyte and the exclusion of the gap in cartilage coverage caused by a protruding central osteophyte in (D). The SBL is indicated as a red line on the femur and a blue line on the tibia on these sagittal slices, respectively.

### Deep learning framework for bone and cartilage shape segmentation

All the cases reviewed by the expert radiologist were used to train a deep neural network to simultaneously extract the bone and cartilage shapes of the tibia and the femur (**Figure S4**). The neural network was based on a well-known deep learning architecture (U-Net) [23], and has the capability to learn different patterns within imaging data. More details can be found in the supplement.

### Measurement of subchondral bone length

We extracted the SBLs on both the tibia and the femur to capture the shape of the cortical bone of the femur or the tibia that is in contact with the cartilage. Briefly, we applied a distance transform on the output of the U-Net model to detect the edge of the bone region in contact with the cartilage (**Figure 1B**). The distance transform measures the distance of each pixel on the bone from its closest pixel on the cartilage. We subsequently skeletonized the detected region to a thickness of one pixel and calculated its arc length. If there were multiple regions detected in the femur or the tibia for a specific 2D sagittal slice, then we skeletonized each region individually and summed the arc lengths of each segment as SBL. For instance, areas denuded of cartilage would produce a gap in the bone length, thus the computed SBL would be the sum of the segments of the cartilage-covered bone. These measurements were confirmed by the radiologist.

The SBL estimates for both the femur (red curved lines in **Figures 1C & 1D**), and the tibia (blue curved lines in **Figures 1C & 1D**), were extracted from each 2D sagittal slice at various locations along the frontal axis (medial to lateral side) of each knee (**Figures 2A, 3A & 4A**). The knees were further stratified by JSN grade within the medial and lateral compartments on the femur and the tibia.

**Figure 2:**
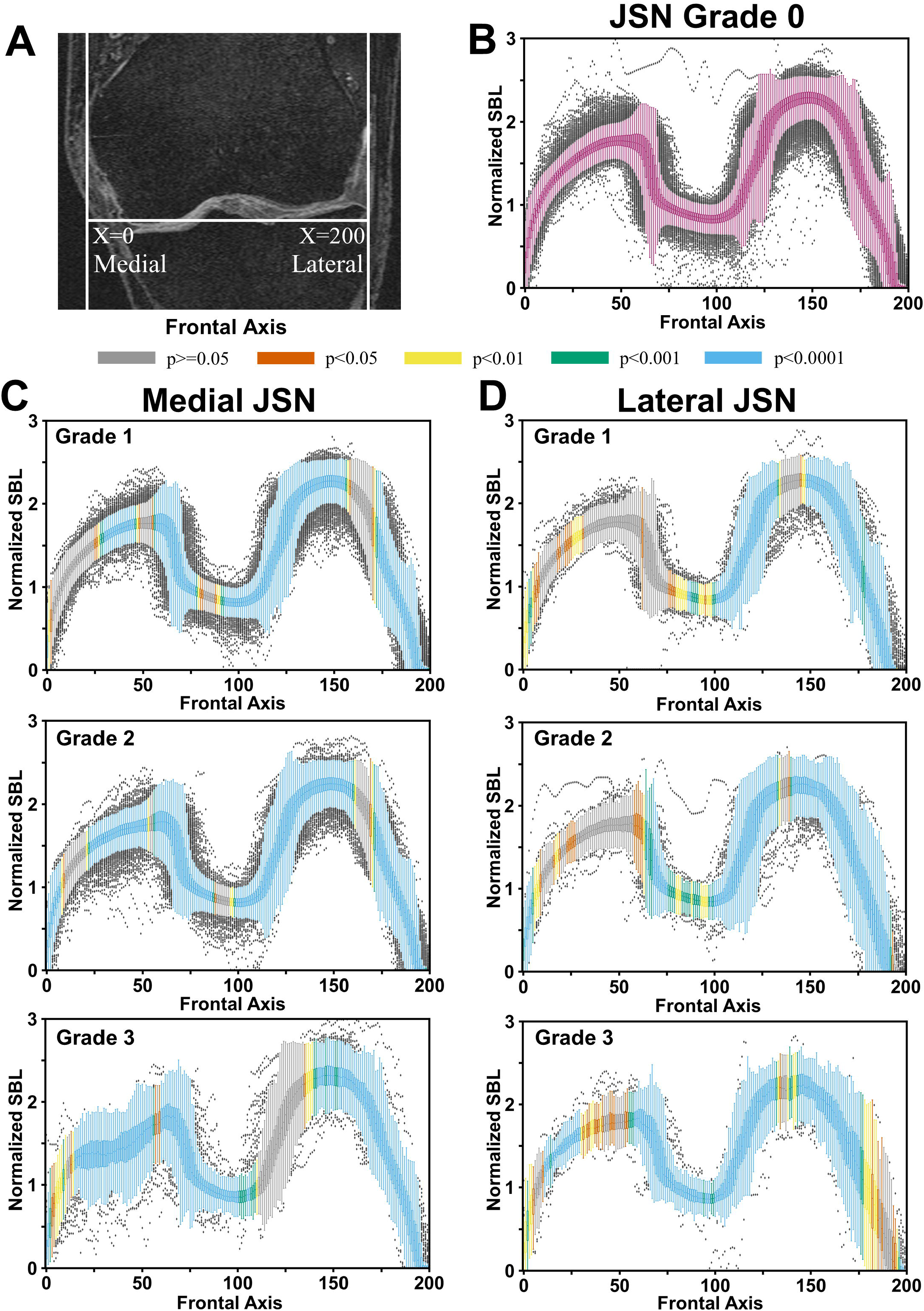
Distribution of subchondral bone lengths on the femur. (A) Placement of the normalized x-axis on the MRI slices. For each location on the x-axis shown in the coronal plane, subchondral bone lengths (SBLs) were computed on the sagittal slices for all the knees. A total of 200 locations were chosen along the frontal axis. (B) Box plots containing the normalized femoral SBLs for JSN=0. (C) Normalized SBL values plotted along the frontal axis for different grades of medial JSN. (D) Normalized SBL values plotted along the frontal axis for different grades of lateral JSN. The SBL normalization was performed by dividing the SBL estimated per slice with the summation of all the femur and tibial SBLs for a given knee. The color coding on the box plots at every location denote the p-value computed using the corresponding location on the JSN=0 plot as the reference.

**Figure 3:**
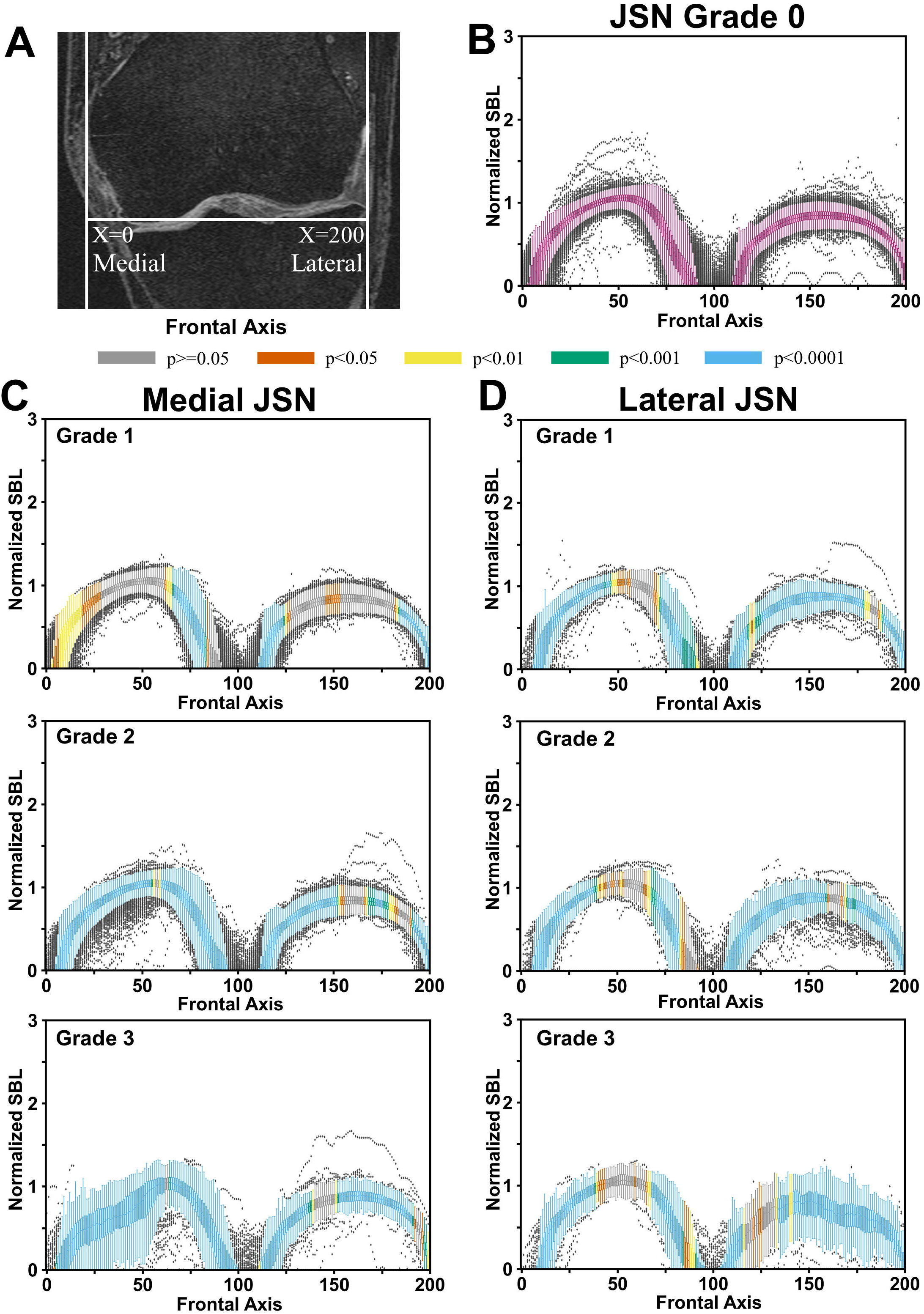
Distribution of subchondral bone lengths on the tibia. (A) Placement of the normalized x-axis on the MRI slices. For each location on the x-axis shown in the coronal plane, subchondral bone lengths (SBLs) were computed on the sagittal slices for all the knees. A total of 200 locations were chosen along the frontal axis. (B) Box plots containing the normalized tibial SBLs for JSN=0. (C) Normalized SBL values plotted along the frontal axis for different grades of medial JSN. (D) Normalized SBL values plotted along the frontal axis for different grades of lateral JSN. The SBL normalization was performed by dividing the SBL estimated per slice with the summation of all the femur and tibial SBLs for a given knee. The color coding on the box plots at every location denote the p-value computed using the corresponding location on the JSN=0 plot as the reference.

**Figure 4:**
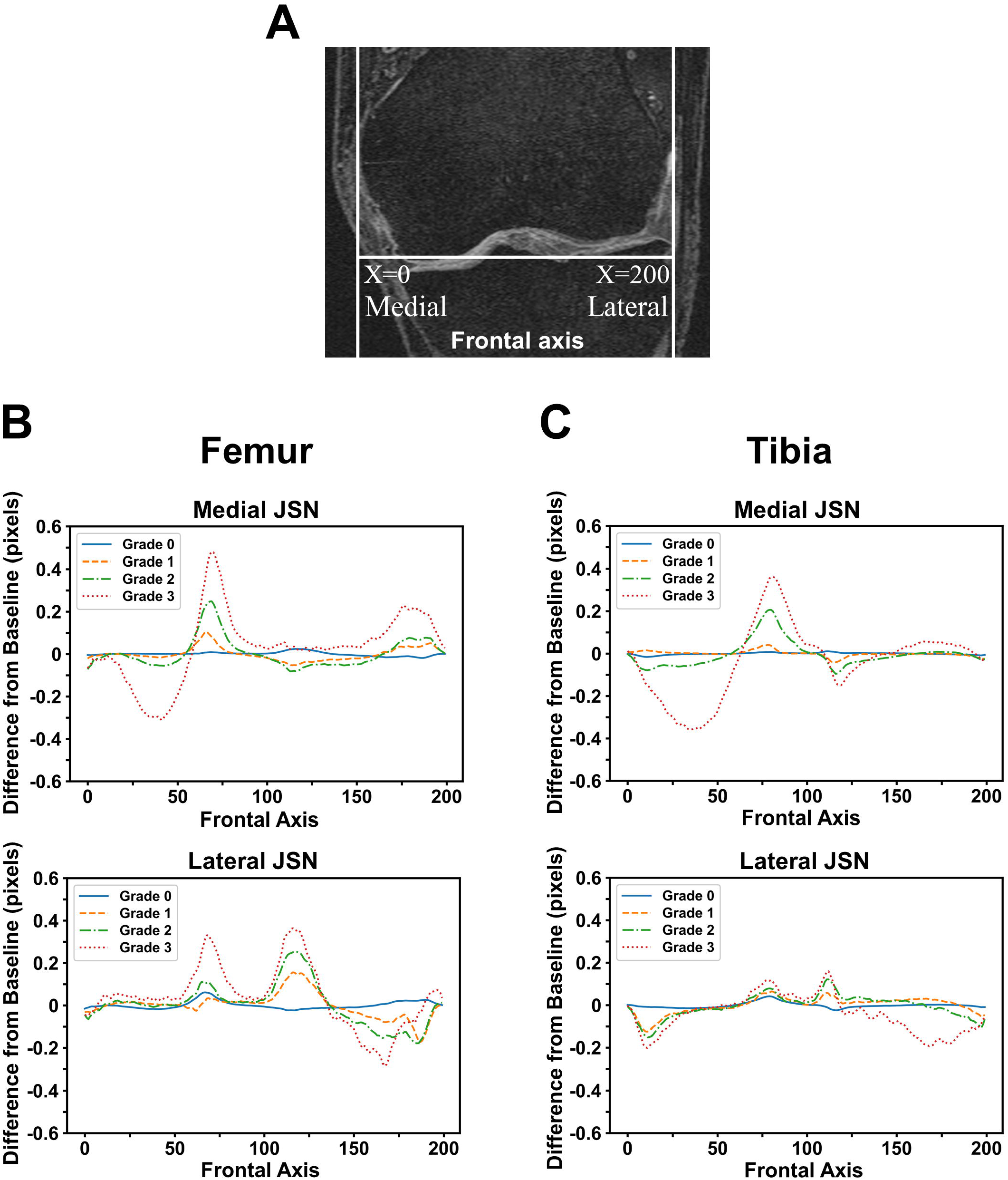
Differences in mean subchondral bone lengths. (A) Placement of the normalized x-axis on the MRI slices. (B) Difference in mean femoral SBLs compared for cases with different grades of medial or lateral JSN. (C) Difference in mean tibial SBLs compared for cases with medial or lateral JSN. A total of 200 locations were chosen along the frontal axis. The units of the mean SBL values plotted on y-axis are the number of MRI pixels separating the mean value of SBL knees with the JSN grade.

### Statistical analysis

The deep neural network used for segmenting the bone shapes was evaluated by computing the Dice coefficient between the predicted masks (generated from the U-Net model), and the bone shapes outlined by the radiologist on the test cases (n=27). Descriptive statistics are presented as mean values and standard deviations. A Student’s t-test was used to compare the mean value of two different groups. A p-value of <0.05 was considered statistically significant. To compare SBLs between knees of various sizes, we took the calculated SBL values from the MRI slices where cartilage was visible and interpolated the values uniformly along the frontal axis (**Figures 2A, 3A & 4A**). For the SBLs taken from the n^th^ location along the frontal axis, we applied a t-test comparing the SBLs of knees with only lateral JSN (374 right and 342 left knees) with knees with no JSN (5128 right and left knees), and knees with only medial JSN (1604 right and 1504 left knees) with knees with no JSN (5128 right and left knees).

We computed mean estimates of femur-specific and tibia-specific SBL measurements at each location on the frontal axis for all the knees with JSN=0. For each knee with JSN>0, we computed difference between the SBL value measured at the n^th^ location and the corresponding mean SBL estimate at that location in knees with JSN=0. We then added the absolute value of these differences to create knee-specific SBL measures and divided them into quartiles. Each group (medial or lateral JSN cases) was further stratified based on clinical outcomes, which included baseline WOMAC pain and disability scores and future total knee replacement. The baseline pain and disability scores were divided by severity, such that scores for pain ≥4 and <8 were moderate and those ≥8 were severe. Similarly, scores for disability ≥20 and <35 were moderate and those ≥35 were severe [11]. The criterion for future knee replacement was the subject having a knee replacement seen on follow-up x-ray at any time in an 8-year follow-up after baseline.

We calculated odds ratios by comparing the odds of each quartile developing the various outcomes, using Quartile 1 as a reference. The odds ratios were calculated using a 2×2 contingency table with binomial outcomes for the pain and disability scores such that one outcome was compared to the remaining outcomes in its corresponding category. For example, the odds of having moderate pain were compared to no pain and severe pain combined. Severe pain was compared with the combination of moderate and no pain. The p-values were calculated using the Fisher exact test. Python scripts are made available on GitHub (https://github.com/vkola-lab/ar2021).

## RESULTS

### Manual versus automated segmentation

Using the deep learning framework, we estimated the bone shapes defining the tibiofemoral joint (**Figure S5**). The Dice coefficients for the femur cartilage, medial and lateral tibia cartilage, on the U-Net model were 0.903, 0.913, and 0.873, respectively. Comparison of the manual annotation and automated segmentation of the femur, the tibia, and the meniscus with Dice coefficients demonstrated the similarity between the predicted and expert-driven segmentations for the U-Net model. The red and blue lines in the figures indicate SBLs connected with the femoral and tibial cartilage, respectively (**Figures 1C & 1D**). In a further comparison between the respective masks generated by the U-Net model (Predicted) and the expert-driven hand annotated images, scatter plots were generated with the measured SBL of each MRI slice of the reference estimate against predicted values (n=27 subjects). Reference estimate versus predicted SBLs of femur generated an R^2^ value of 0.922 (**Figure S6A**). Much of the discrepancy between the reference estimate from the radiologist versus predicted SBL values occurred at the extreme medial and lateral sagittal slices (SBL length<50 pixels). This pattern was consistent with the findings in tibial SBL (R^2^=0.902) (**Figure S6B**), with higher discrepancies seen at SBLs of less than 25 pixels. These correspond to the lateral-most and medial-most slices of the knees.

### Subchondral bone length in relation to joint space narrowing

Our analysis showed that most SBL values from knees with JSN >0 were significantly different in length than knees with no JSN for both the femur and the tibia (**Figures 2B-2D & 3B-3D**). This was true for knees with both medial and lateral JSN. In cases of medial JSN, on the femur we observed significant differences in the SBL values for most of the medial-central, central and lateral-most regions (**Figures 2C**). In the cases of lateral JSN, the SBL values were significantly different from JSN=0 knees in most of the lateral regions of the femur (**Figure 2D**), except for a few lateral-most regions for JSN=3. For cases with lateral JSN, statistically significant differences were not observed in most of the medial-central regions of the femur, regardless of the JSN grade (**Figure 2D**).

In cases of medial JSN on the tibia, we observed statistically significant differences in the medial and central regions for JSN>1 but only in the central regions for JSN=1 (**Figure 3C**). No significant differences were observed in the medial-central and the lateral-central regions for JSN=1 (**Figure 3C**). In cases of lateral JSN for the tibia, differences were observed in the medial-most and lateral-most regions for all JSN grades (**Figure 3D**). The variability in the SBL estimates seemed highest for medial-central and medial-most regions when there was medial JSN, and in the lateral-central and lateral-most regions when there was lateral JSN for JSN=3 in the tibia (**Figures 3C & 3D**).

When we further examined the mean differences along the frontal axis for the femur and the tibia, the variability in the SBL distribution increased with JSN grade. Notably, cases with JSN=3 had mean SBL values with higher peaks and troughs than JSN<3, regardless of the location of JSN (**Figure 4**). For medial JSN=3 in the femur and the tibia, the trough was in the middle of the medial compartment whereas for lateral JSN=3, it was in the far lateral part of the lateral compartment. SBL values closer to the middle of the knee in the frontal axis showed an increase in SBL compared with JSN=0 (**Figures 4B** & **4C**). While reductions in SBLs were seen in some locations within the affected compartments, regions near the middle of the knee often showed increased SBL lengths compared with knees that had JSN=0.

### Subchondral bone length in relation to outcomes

In cases with medial JSN, the odds ratios (OR) for moderate pain and disability was highest for knees in the highest quartile of SBL (**Tables S1 & 2**). Among knees with medial JSN >0, the odds of severe pain at baseline increased with increasing quartile of SBL, increasing about 4-fold (OR = 4.09 (95% CI: 2.88, 5.79)) for knees in the highest quartile. The ORs for severe disability showed a similar pattern. Among these knees, the odds of future TKR increased also with increasing SBL, rising to 5.68 (95% CI: 3.90, 8.27) for knees in the highest quartile. For knees with lateral JSN (**Table 2**), the pattern was similar, with increasing odds of pain and disability and increased odds of TKR for knees with higher SBLs.

**Table 2:** Association of SBL with various outcomes. Odds ratios (OR) and 95% confidence intervals for SBL quartiles among JSN grades ≥1, compared with the lowest SBL quartile, for various outcomes.

| Condition | Outcome | Quartile 2 (OR [CI]) | Quartile 3 (OR [CI]) | Quartile 4 (OR [CI]) |
| --- | --- | --- | --- | --- |
| JSN Medial (≥1) | Moderate Pain (≥4, <8) | 0.90 [0.69, 1.17] | 1.31* [1.02, 1.68] | 1.60*** [1.26, 2.04] |
|  | Severe Pain (≥8) | 2.49*** [1.72, 3.58] | 2.24*** [1.55, 3.24] | 4.09*** [2.88, 5.79] |
|  | Moderate Disability (≥20, <35) | 1.10 [0.81, 1.50] | 1.51** [1.13, 2.02] | 1.86*** [1.40, 2.48] |
|  | Severe Disability (≥35) | 2.90*** [1.60, 5.28] | 2.47** [1.34, 4.55] | 4.33*** [2.44, 7.68] |
|  | Future TKR | 1.45 [0.93, 2.24] | 2.53*** [1.69, 3.78] | 5.68*** [3.90, 8.27] |
| JSN Lateral (≥1) | Moderate Pain (≥4, <8) | 1.29 [0.79, 2.11] | 1.29 [0.79, 2.11] | 1.49 [0.91, 2.42] |
|  | Severe Pain (≥8) | 1.40 [0.72, 2.73] | 1.92 [1.02, 3.62] | 3.11*** [1.70, 5.68] |
|  | Moderate Disability (≥20, <35) | 1.17 [0.62, 2.22] | 1.86 [1.03, 3.38] | 2.51** [1.41, 4.48] |
|  | Severe Disability (≥35) | 2.28 [0.78, 6.70] | 2.06 [0.69, 6.15] | 3.89** [1.41, 10.72] |
|  | Future TKR | 2.36* [1.15, 4.85] | 2.15 [1.04, 4.46] | 7.19*** [3.71, 13.95] |
| *p<0.05, **p<0.01, ***p<0.001 |  |  |  |  |

## DISCUSSION

In this work, we introduced a new MRI-derived bone measure defined as subchondral bone length (SBL) by leveraging a deep learning-based image segmentation framework and expert-driven annotations of structures comprising the knee joint. SBL depicts both tibiofemoral articular cartilage morphology and bone shape, as it reflects the cartilage-covered articular area translated to length by excluding osteophytes and denuded areas. As accurate manual segmentation of MRI studies can be highly labor-intensive, there have been several efforts to develop computational methods for automating the segmentation and the measurement of knee structures with specific focus on the cartilage and the meniscus [24-31], and other structures of the knee joint [25, 32-34]. We performed additional analyses to show that SBL values at different locations of the knee vary in magnitude as a function of JSN grade in both the femur and the tibia. Finally, we showed that high mean SBL values identify knees with high risk of pain and disability and with an increased risk of later TKR.

Imaging biomarkers visualized solely on MRI, such as subchondral changes, cartilage volume, and bone marrow lesions, have been found to have better correlation with symptomatic presentation in patients compared to findings on x-rays like KL grade [35]. If we measure subchondral changes on MRI at the slice level, then we can expand the range of analyses by considering how specific regions of the subchondral bone correlate with the progression and outcomes of knee OA. Our work is unique in focusing on SBL as a shape measure and provides novel insights into where bone length changes with increasing disease severity. There are two factors affecting this planar bone length measure, cartilage loss which creates gaps in bone length and shortens it, and flattening of the bone which can have the opposite effect, causing an increase in length. Since SBL shows shortening only with complete local loss of cartilage and not cartilage thinning, the dominant effect of disease is to cause SBL lengthening. Because of flattening, bone lengthens in affected compartments even with removal of osteophytes [15]. We observed that mean SBL values displayed an interesting dynamic on the knees with JSN>0 versus those with JSN=0, suggesting that cartilage loss and increased bone length both drive this measure. Consistent with studies of where cartilage loss occurs with advancing OA [36], the mean SBL was shorter in the peripheral sub-regions of the knee. Also, mean SBL values followed a trend where the peaks occurred consistently at similar locations along the frontal axis, where the magnitude of these peaks was a function of the JSN grade. We would expect with increasing cartilage loss that SBL would decrease but also increase due to bone flattening. Since the odds of TKR and other outcomes increased with increasing SBL, it implies that SBL changes were predominantly influenced by bone flattening. The same reasoning would apply to decreasing joint space widening based on OARSI scales such that SBL would be expected to increase in those cases as well. Nevertheless, understanding region-specific subchondral bone lengthening with developing disease may provide insights into how bone response to loading affects disease in different regions of the knee.

SBL shows promise as an imaging measure for analyzing OA. It allows for feasible statistical analysis of both the amount of exposed bone from cartilage loss and the remodeling of the cortical envelope that occurs as part of the disease process. By utilizing the length instead of the subchondral surface area, it becomes straightforward to derive insights based on slice location into common regions of joint degeneration in different presentations of knee OA. Because SBL was derived by excluding marginal osteophytes, this shape measure is distinct from other published measures of bone shape dependent on the crust of the osteophytes around the edges of the articular cartilage [8]. Marginal osteophytes do not appear to be related to joint pain. Further, atrophic and hypertrophic forms of disease have similar levels of cartilage loss, and studies suggest that marginal osteophytes are often not in the same compartment as cartilage loss and can even occur with no loss [14]. Lastly, molecular stimuli of osteophyte growth often have no measurable effect on cartilage loss [37, 38]. SBL measurement may become all the more relevant because this feature could facilitate future studies related to quantifying the effects of subchondral bone changes on transarticular loading patterns across the knee and provide a better understanding of OA progression.

From the OAI database, we acquired demographic data that allowed for analysis of correlations between JSN and gender, age, race, and BMI. Using our U-Net model, we could also apply automated segmentation to find correlations between specific knee structure changes in OA with other biomarkers. These correlations can be used to determine risk factors and individual susceptibility to develop knee OA. Also, it would be possible to visualize biomarker changes in relation to bone structure over time. Since the entire process is automated, these analyses can be done in an efficient fashion.

One of the main challenges in applying deep learning to perform automated segmentation of medical imaging is the lack of high-volume expert-annotated data. To circumvent this, we adapted an efficient strategy by utilizing the DRLSE method in combination with human verification to generate a large volume of bone annotations from the MRIs of 61 knees to train the U-Net model. The automated segmentations of bone and cartilage from the U-Net were subsequently compared against the annotation of an expert radiologist, which showed high accuracy in terms of the Dice coefficient.

There are a few limitations to our study. Our deep learning framework is based on a 2D U-Net architecture, and recent studies have attempted to employ more advanced frameworks involving 3D neural network architectures for image segmentation tasks [25]. However, the selection of the 2D architecture was sufficient to develop SBL as a novel 2D shape measure. Also, more studies are needed to understand the distributions of mean SBL values for various grades of disease, and evaluate whether this effect is due to cartilage loss or flattening of the bone or some combination of both. There are some areas of the knee where SBL measurement is likely to be challenging. These areas correspond to subchondral sagittal slices at the medial-most and lateral-most aspects of the knee. It is at this region that the defining border between cartilage and bone becomes unclear, thus complicating the segmentation process for radiologists as well as our automated segmentation model. Furthermore, these portions of the knee are common locations for the development of osteophytes, leading to irregular SBL that may lead to variability and poor correlation with scoring systems that utilize osteophytes as a factor.

In conclusion, our study demonstrates the ability for a deep learning framework to learn from expert-driven annotations and extend it to generating a quantitative understanding of the knee joint structures across a large cohort. Such extensions have the potential to generate statistically significant representations of knee joint shapes without having to manually process imaging data from all the cases. As an example, we identified SBL as a potentially useful measure of the subchondral bone morphology within the knee joint and showed that it varies with JSN grade. Our findings can assist in a more detailed quantification of shape-specific risk factors of knee OA.

## Data Availability

Python scripts are made available on GitHub (https://github.com/vkola-lab/ar2021).

## SUPPLEMENT FIGURES

**Figure S1:**
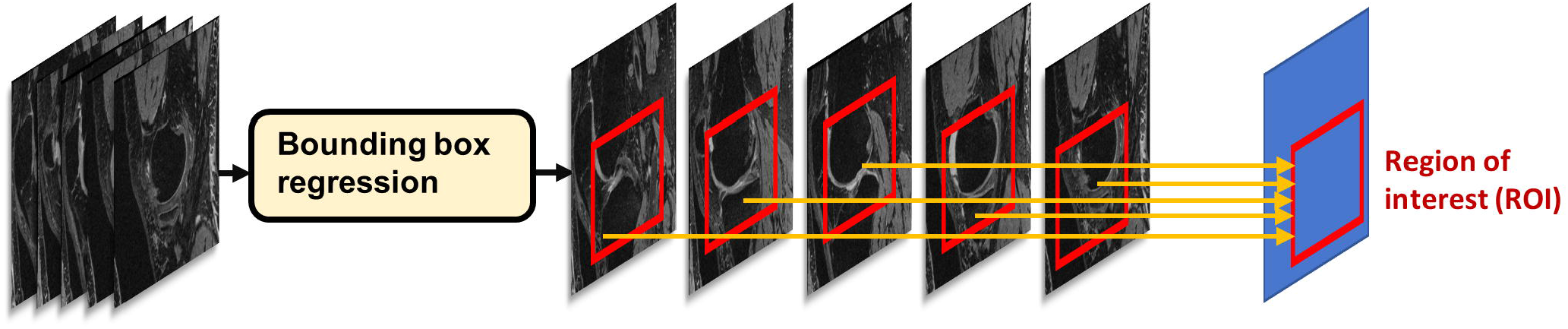
A fast, region-based convolutional neural network (Fast R-CNN) to identify the location of the knee joint in two-dimensional sagittal slices.

**Figure S2:**
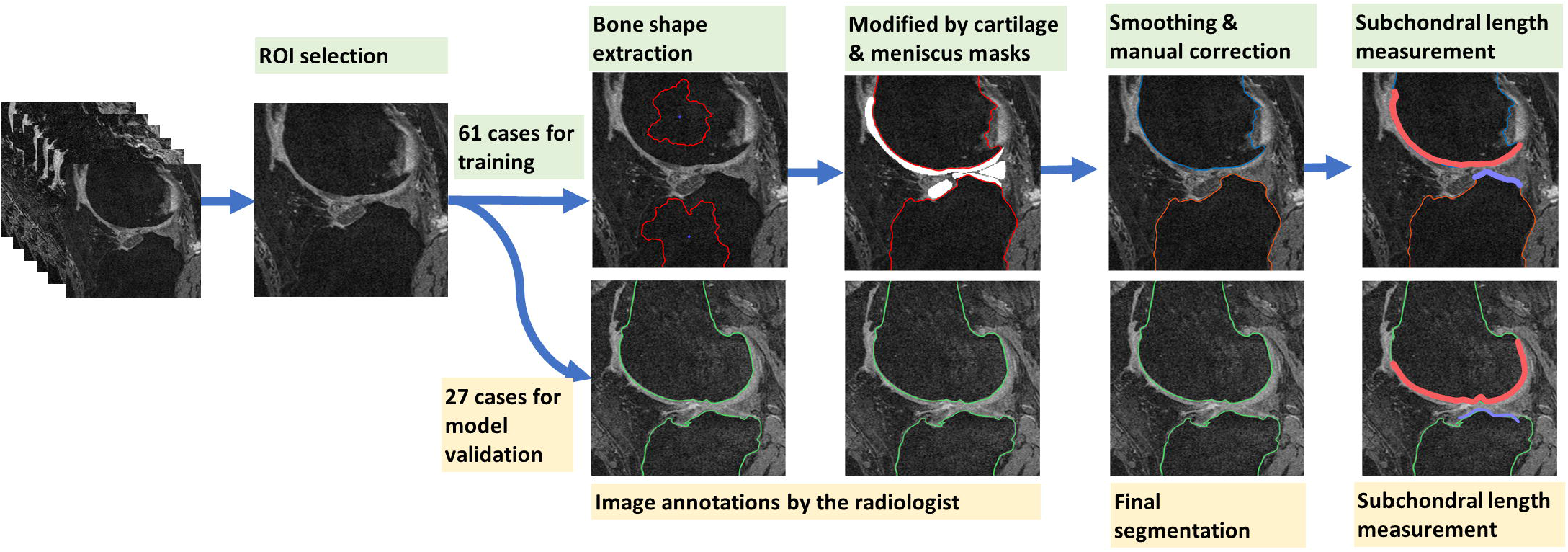
The outputs of the R-CNN model were used to generate annotations of the femur and tibia on 88 cases. A portion of these cases (n=61) were used for training a two-dimensional U-Net, and the remaining cases (n=27) were used for validation. All the validation cases were reviewed by the radiologist. The SBLs are indicated as red line on the femur and blue line on the tibia, respectively.

**Figure S3:**
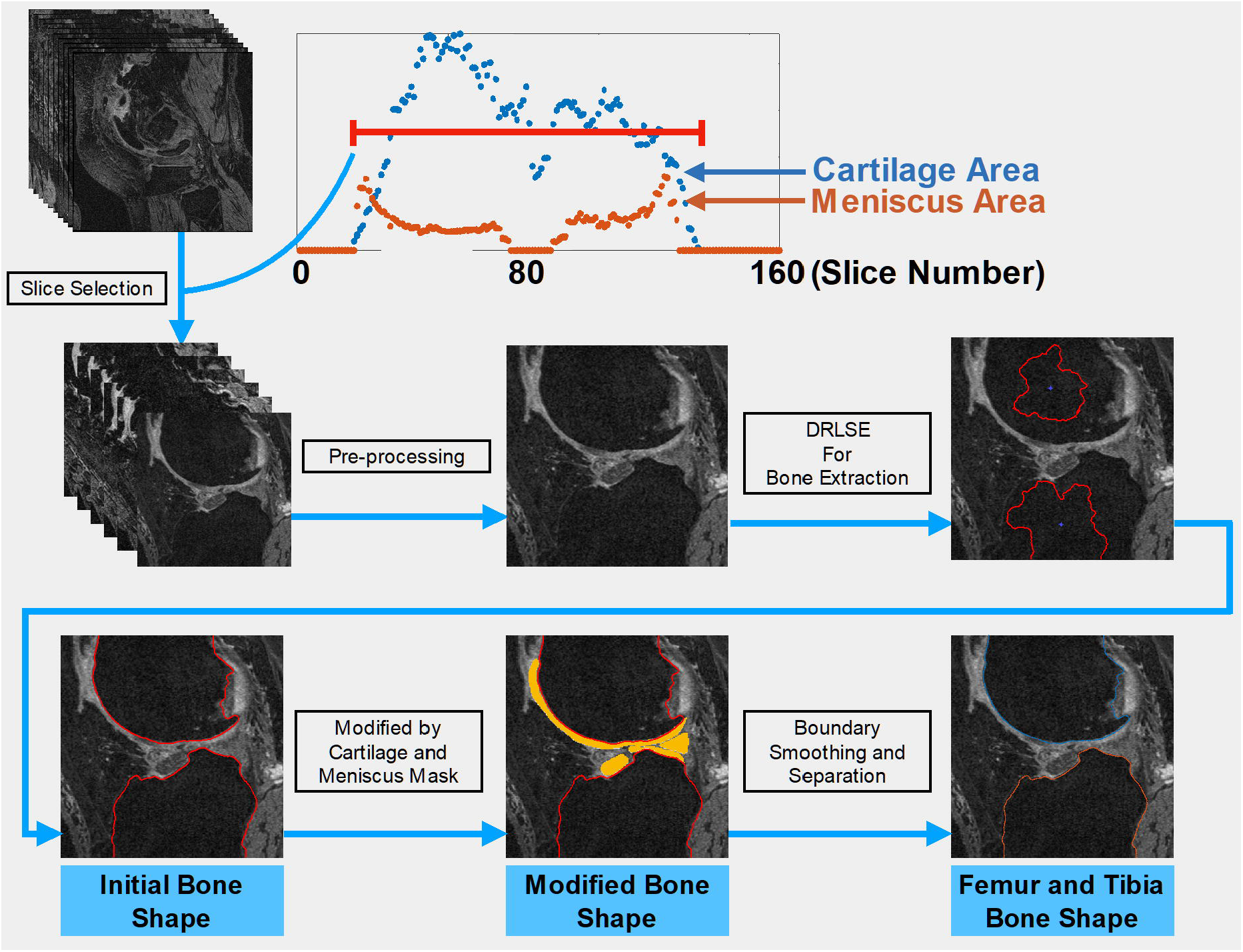
Generation of masks for the subchondral regions using Distance Regularized Level Set Evolution (DRLSE). These masks were used as training data to construct the U-Net model.

**Figure S4:**
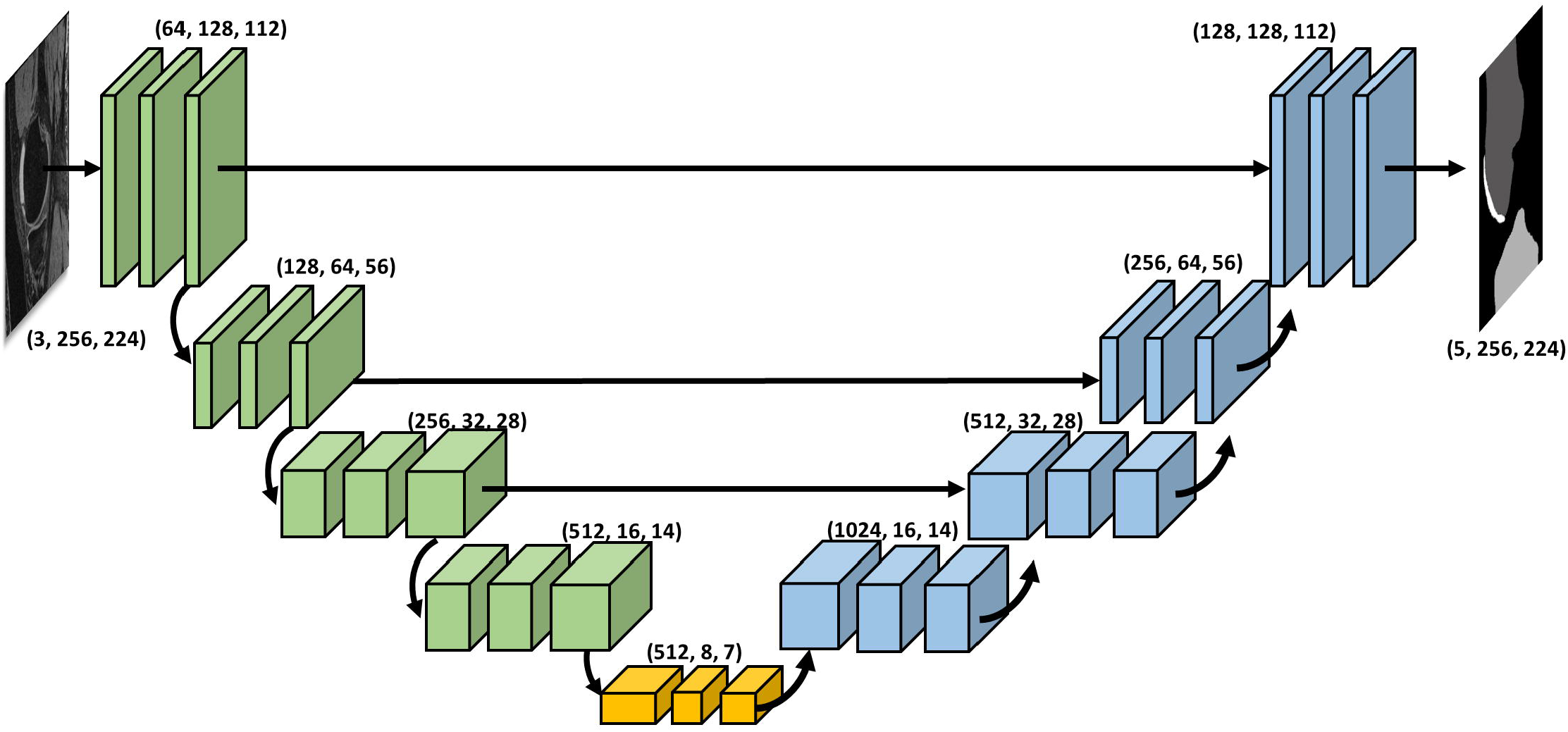
Two-dimensional U-Net architecture used to segment tibiofemoral subchondral bone and cartilages from 2D MRI slices.

**Figure S5:**
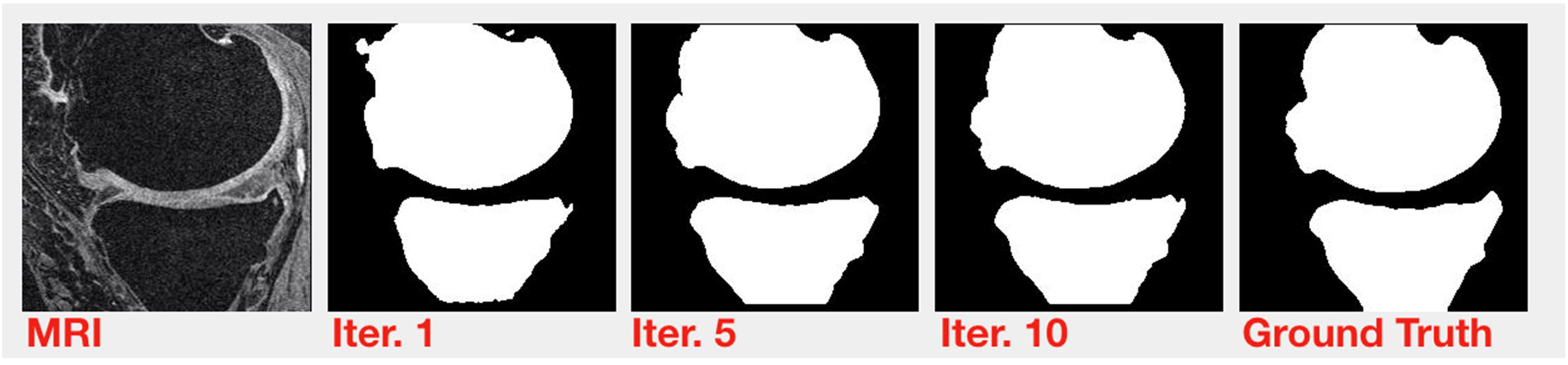
Evolution of the femur and bone segmentation process over several iterations. The original MRI slice (sagittal view) along with the annotation performed by the radiologist (reference estimate), are also shown.

**Figure S6:**
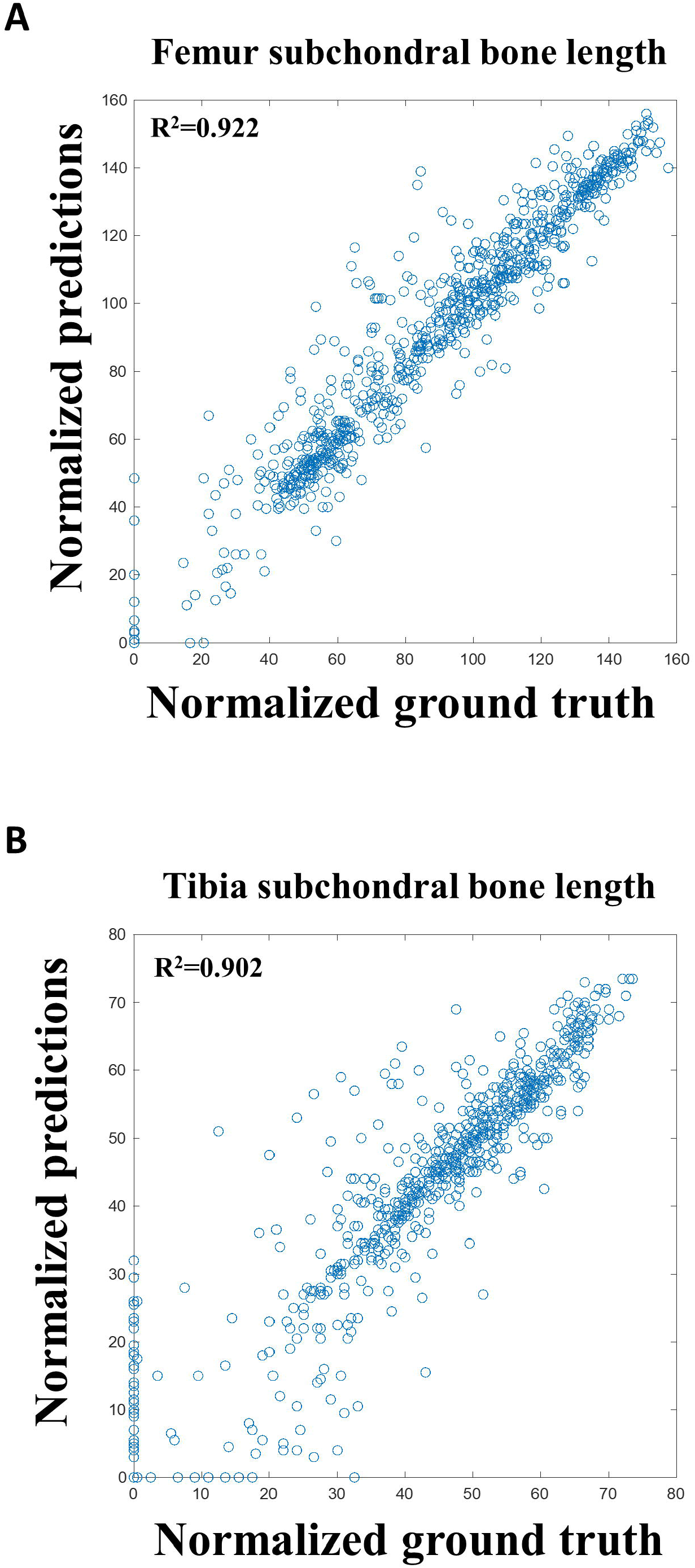
Model predictions versus measured sub-chondral bone lengths for (A) the femur and (B) the tibia.

**Table S1:** Odds ratio descriptive statistics of SBL for various groups. Details including mean, standard deviation, and median SBL values along with the total number of cases are shown.

| Odds Ratio Descriptive Statistics of SBL Difference (Mean, Standard Deviation, Median, Total) |  |  |  |  |  |
| --- | --- | --- | --- | --- | --- |
| Group |  | Q1 | Q2 | Q3 | Q4 |
| JSN Medial >=1 |  | 30.59, 3.11, 31.15<br>n=777 | 38.18, 1.95, 38.23<br>n=777 | 46.15, 2.88, 45.91<br>n=777 | 67.95, 19.17, 62.35<br>n=777 |
|  | Moderate Pain | 30.66, 3.16, 31.20<br>n=142 | 38.06, 1.98, 38.03<br>n=130 | 46.42, 2.86, 46.11<br>n=176 | 70.62, 22.00, 64.47<br>n=205 |
|  | Severe Pain | 31.36, 2.85, 32.12<br>n=45 | 38.30, 1.97, 38.34<br>n=103 | 46.32, 2.91, 46.15<br>n=94 | 71.78, 21.86, 64.86<br>n=156 |
|  | Moderate Disability | 30.91, 3.07, 31.89<br>n=89 | 38.03, 1.88, 38.05<br>n=97 | 46.47, 2.90, 46.10<br>n=127 | 72.98, 26.07, 65.49<br>n=151 |
|  | Severe Disability | 32.20, 2.37, 32.70<br>n=15 | 38.27, 2.14, 38.32<br>n=42 | 46.67, 3.03, 46.54<br>n=36 | 70.66, 19.13, 64.57<br>n=61 |
|  | Future KR | 31.83, 2.64, 32.37<br>n=36 | 38.52, 1.97, 38.89<br>n=51 | 46.72, 3.06, 46.73<br>n=85 | 76.57, 25.13, 69.57<br>n=168 |
| JSN Lateral >=1 |  | 34.28, 3.69, 34.80<br>n=179 | 44.31, 2.99, 44.25<br>n=179 | 55.70, 3.66, 55.75<br>n=179 | 79.84, 22.66, 73.49<br>n=179 |
|  | Moderate Pain | 34.04, 3.62, 34.70<br>n=37 | 44.47, 3.13, 44.25<br>n=45 | 55.86, 3.52, 56.37<br>n=45 | 81.25, 18.01, 73.82<br>n=50 |
|  | Severe Pain | 34.98, 3.99, 35.99<br>n=17 | 44.41, 3.31, 44.53<br>n=23 | 54.63, 2.89, 54.86<br>n=30 | 85.39, 27.53, 78.44<br>n=44 |
|  | Moderate Disability | 34.38, 3.36, 34.75<br>n=20 | 44.11, 3.43, 43.57<br>n=23 | 55.55, 3.37, 55.38<br>n=34 | 85.28, 29.84, 74.43<br>n=43 |
|  | Severe Disability | 35.30, 5.24, 37.02<br>n=5 | 43.46, 2.95, 43.19<br>n=11 | 54.41, 4.04, 54.51<br>n=10 | 83.56, 12.32, 86.24<br>n=18 |
|  | Future KR | 35.03, 3.49, 36.67<br>n=12 | 44.49, 3.52, 44.38<br>n=26 | 56.18, 3.63, 55.85<br>n=24 | 79.68, 13.85, 76.48<br>n=61 |

